# Cost-effectiveness and cost-utility of antenatal sexually transmitted infection screening to reduce preterm birth and low birthweight in South Africa

**DOI:** 10.64898/2026.08.08.26360003

**Authors:** Elise Smith, Chibuzor M Babalola, Andrew Medina-Marino, Mandisa Mdingi, Freedom Mukomana, Nicola Low, Amarech Obse, Remco P.H. Peters, Jeffrey D Klausner, Susan Cleary, Edina Sinanovic

## Abstract

**Background:** Curable sexually transmitted infections (STIs) are associated with adverse birth outcomes, yet little cost-effectiveness evidence guides antenatal STI screening policy in high-burden settings. We conducted a cost-effectiveness and cost-utility analysis of the Philani Ndiphile trial in South Africa, comparing One-Time and Two-Time antenatal screening for *C. trachomatis, N. gonorrhoeae,* and *T. vaginalis* with standard syndromic management.

**Methods:** A decision-analytic model from the provider perspective simulated costs and outcomes for pregnant women and infants. Costs included diagnostics, treatment, and neonatal hospitalisation for a primary composite outcome of preterm birth and/or low birthweight and its components (secondary trial outcomes). Modelled outcomes included incremental cost (US$) per composite (preterm birth and/or low birthweight) case, per component case and per disability-adjusted life year (DALY) averted. The analysis captured infant outcomes in the first year. Univariate and probabilistic sensitivity analyses were conducted to assess parameter uncertainty and robustness of results.

**Results:** Screening for *C. trachomatis, N. gonorrhoeae,* and *T. vaginalis* twice during pregnancy was not cost-effective for preventing the primary composite outcome, nor for preventing low birthweight alone. However, two-time screening was cost-saving for preventing preterm birth alone, averting more DALYs and thereby yielding better health outcomes while reducing healthcare costs compared with syndromic management. One-time screening was not cost-effective for preventing any outcome.

**Conclusions:** Repeat antenatal screening for *C. trachomatis, N. gonorrhoeae*, *and T. vaginalis* has the potential to prevent preterm births while reducing healthcare costs in high-burden settings. These findings support further research to confirm the clinical effectiveness of repeat screening and to evaluate longer-term health and economic impacts.

## BACKGROUND

The prevalence of curable sexually transmitted infections (STIs) such as *C. trachomatis, N. gonorrhoeae,* and *T. vaginalis* is highest in low- and middle-income countries, and has been reported to exceed 30% among pregnant women in some settings.[1, 2] STIs have been associated with poor birth outcomes in observational studies, including preterm birth, low birthweight, and congenital infections.[3–5] Prematurity is the single largest contributor to neonatal deaths globally, accounting for 37%.[6] In South Africa, the World Health Organization (WHO) estimated preterm birth and low birthweight prevalence rates of 13% and 17% respectively in 2020.[7]

In resource-limited settings, WHO recommends syndromic management of STIs. This low-cost approach treats symptom-based syndromes (e.g., vaginal discharge, lower abdominal pain), with combination antibiotic regimens, but fails to address asymptomatic infections.[8–11] The Southern African HIV Clinicians Society (2022) recommends antenatal STI screening at booking and in the third trimester[12] and WHO (2025) issued conditional recommendations for antenatal *C. trachomatis* and *N. gonorrhoeae* screening in high-burden settings.[13] However, these recommendations are not based on rigorous randomised controlled trials, and few countries have adopted national screening policies, highlighting a gap between guidance and implementation.[14]

Advances in point-of-care (PoC) molecular diagnostics have improved the feasibility and accuracy of antenatal STI screening and treatment,[11] but evidence on the effect of STI screening and treatment during pregnancy on preterm birth or low birthweight remains uncertain and largely observational.[15–17] In the Philani Ndiphile trial, molecular PoC testing for *C. trachomatis, N. gonorrhoeae,* and *T. vaginalis* at the first antenatal care visit and again during the third trimester did not significantly reduce the primary composite outcome of preterm birth and/or low birthweight compared with syndromic management.[18, 19] However, in pre-specified secondary analyses, two-time screening reduced preterm birth, but not low birthweight.

Evidence regarding the costs and cost-effectiveness of PoC testing and treatment for STIs (beyond syphilis) during pregnancy remains limited,[16, 20] with high molecular diagnostic costs remaining a barrier in low-resource settings.[21, 22] This study used data from the Philani Ndiphile trial to model the provider perspective cost-effectiveness and cost-utility of two screening strategies for *C. trachomatis, N. gonorrhoeae,* and *T. vaginalis* versus syndromic management to inform policy recommendations.

## METHODS

### Trial design and procedures

This economic evaluation was designed and implemented alongside the Philani Ndiphile three-arm randomised controlled trial in Eastern Cape Province, South Africa.[18] Pregnant women aged ≥18 years attending their first antenatal care (ANC) visit before 27 weeks’ gestation were enrolled between March 2021 and May 2024. Participants in the two intervention groups (Arms 1 and 2) received PoC molecular testing using GeneXpert® CT/NG and GeneXpert® TV (Cepheid, Sunnydale, California, United States of America) with immediate treatment for detected infections at their first ANC visit. In Arm 1, included a test-of-cure approximately 3-5 weeks after treatment (‘One-Time Screening’), whereas Arm 2 included repeat testing between 30 and 34 weeks’ gestation (‘Two-Time Screening’).[18] Participants assigned to Arm 3 (control) received routine syndromic management for abnormal vaginal discharge with a combination antibiotic regimen per national guidelines.[23] The primary outcome of the trial was a composite of preterm birth (<37 weeks’ gestation) and/or low birthweight (<2500 g), with pre-specified secondary analyses of each outcome separately.[18]

### Decision analytic model design and approach

We conducted cost-effectiveness and cost-utility analyses, from a provider perspective, comparing the two screening strategies with standard care using a static decision-analytic model in Microsoft Excel (Figure 1). Consolidated Health Economic Evaluation Reporting Standards reporting is followed.[24]

**Figure 1:**
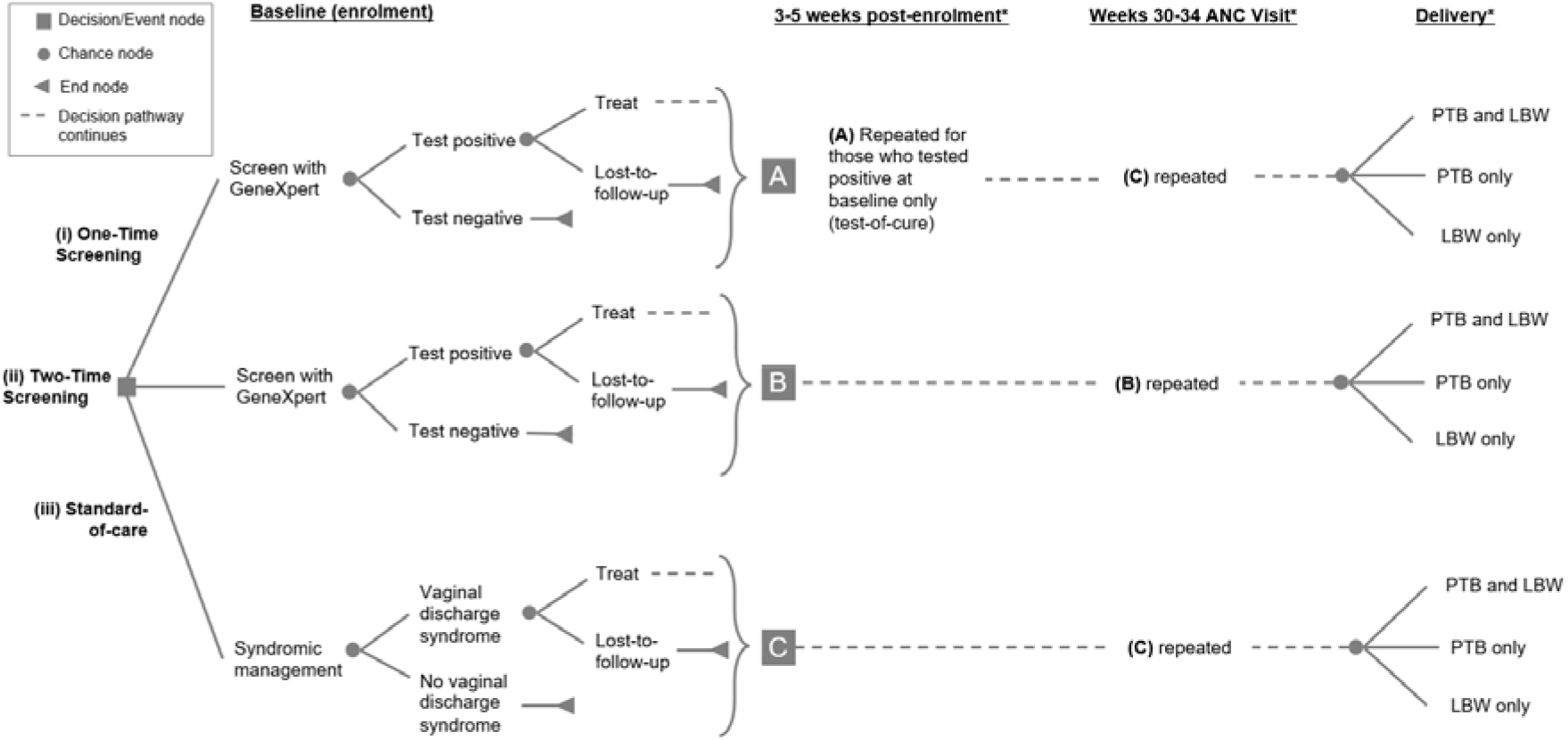
Decision tree model *Lost-to-follow-up between baseline, 3 weeks post-enrolment, week 30-34 ANC visit and delivery not illustrated here; ANC = antenatal care; PTB = preterm birth; LBW = low birthweight

Costs are reported in 2024 United States Dollars (USD; $) using an average exchange rate from 1 January 2024 to 31 December 2024 ($1 USD = R18.55 South African Rand[25]) and inflated, where required, using published consumer price inflation indices.[26] Outcomes included cases averted of the primary composite (preterm birth and/or low birthweight) and secondary component (preterm birth alone; low birthweight alone) outcomes, and modelled disability-adjusted life years (DALYs). Years of life lost due to deaths during the first year of life is discounted at 5%, in line with country guidelines.[27] Trial-based effect sizes and inputs were modelled with uncertainty analyses for all outcomes (Table 1). Cost-effectiveness and cost-utility analyses were conducted regardless of the statistical significance of trial outcomes, consistent with best practice to assess costs and outcomes under uncertainty.[28]

**Table 1:**
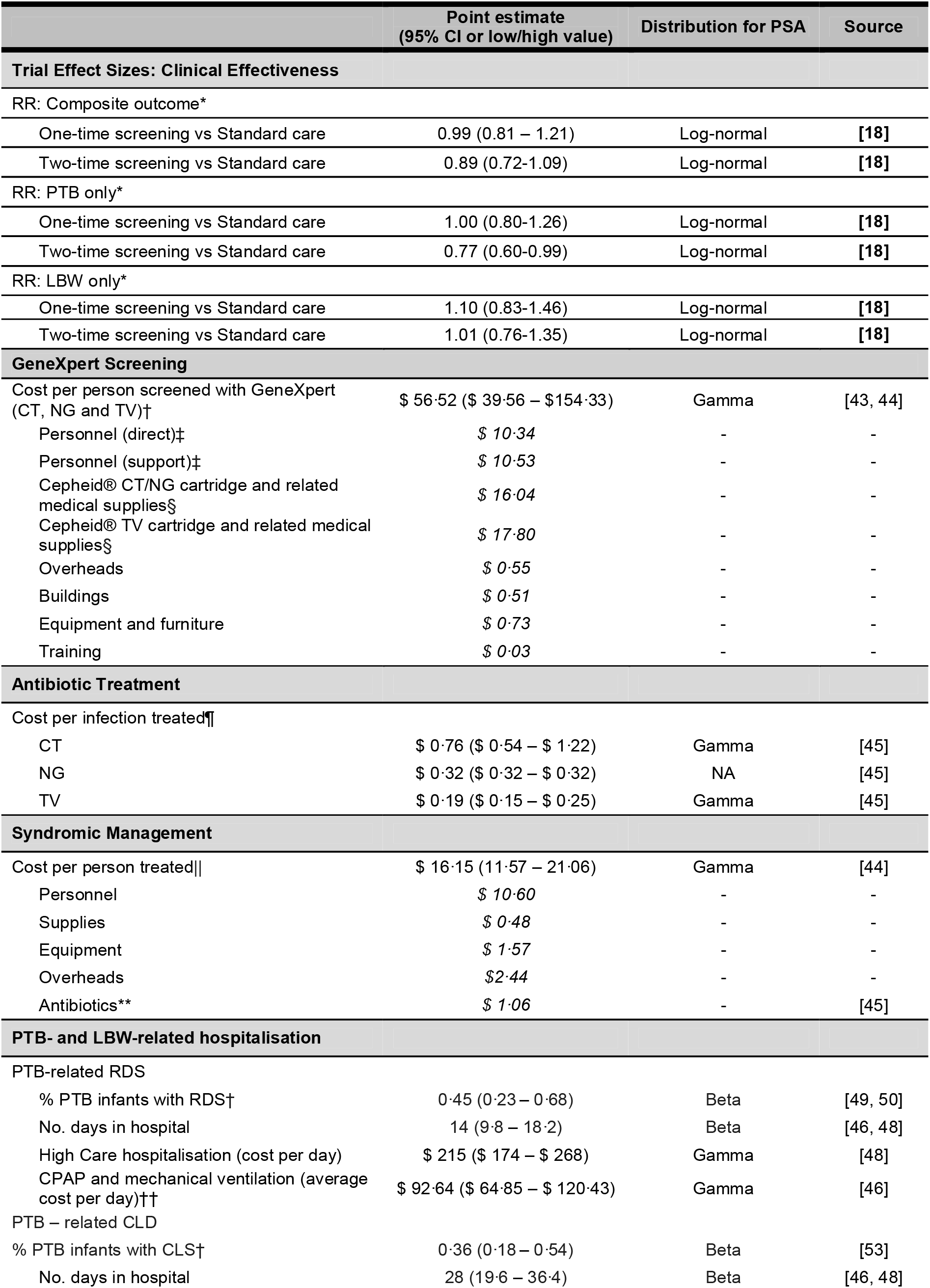
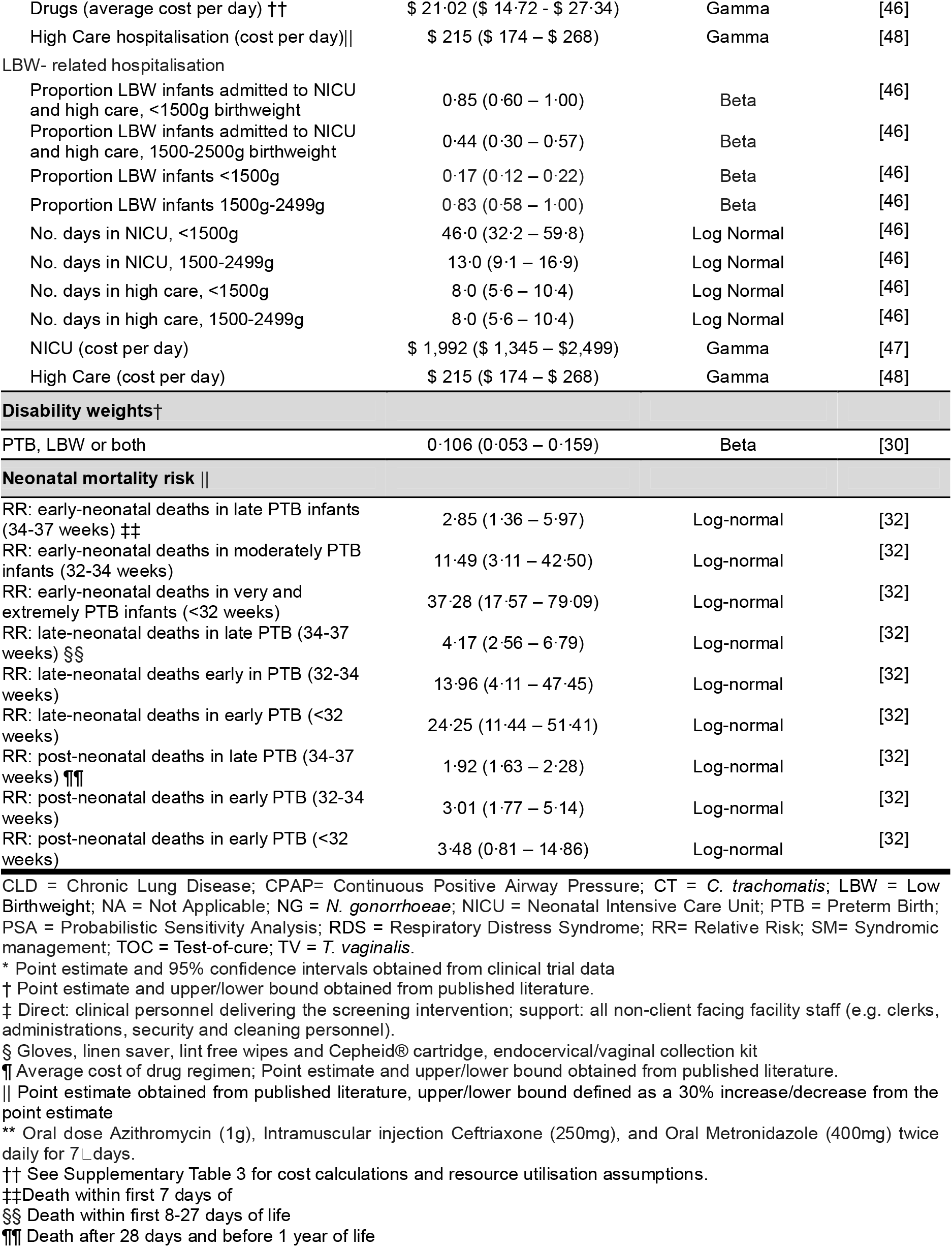
Key model parameters and distributions.

Cost-effectiveness compared intervention and standard-care costs and outcomes, while cost-utility incorporated intervention costs, modelled first-year treatments costs and infant DALYs. Limited long-term data on disability and costs for preterm or low birthweight infants restricted the analysis to the first year of life.

### Health outcomes and probabilities

DALYs were estimated following Fox-Rushby and Hanson[29] without age weighting. [30] Total DALYs per arm comprise years of life lost (YLL) and years lived with disability (YLD) attributable to preterm birth, low birthweight or both.

YLL included deaths within the first year of life (early, late and post-neonatal) attributable to preterm birth, low birthweight or both. Preterm birth-related mortality was calculated using published all-cause mortality rates,[31] including preterm birth mortality rates for each preterm birth category: extremely and very preterm (<32 weeks), moderately preterm (32 to <34 weeks), late preterm (34 to <37 weeks). Preterm birth proportions by category and trial arm are presented in Supplementary Table 1. The relative risk (RR) associated with each preterm birth category in comparison to term birth was also calculated. YLL were calculated by multiplying the country-specific life expectancy with the calculated number of deaths[31–33] (Supplementary Table 2) and discounted at 5%.[27] Owing to limited data, the low birthweight mortality risk was assumed the same as for preterm birth,[34] based on the 2021 Global Burden of Disease Study.

For low birthweight, we used a composite disability weight for low birthweight and its sequelae (0.106)[30, 33, 35–40] We assumed the same disability weight for preterm birth, as previously applied by Price and colleagues[36] (Table 1). YLDs were estimated by applying disability weights to live births, adjusted for the proportion of the first year survived (Supplementary Table 1). Infants born both preterm and with low birthweight were conservatively assumed to incur disability only from preterm birth.[39, 41] We assumed disability weights of zero for losses due to stillbirth and miscarriages. Additional model parameters are detailed in Supplementary Table 2.

### Screening and intervention costs

Costs included antenatal screening intervention and associated treatment, syndromic management, and management of preterm birth and low birthweight complications during the first year of life.

GeneXpert testing costs were estimated using an ingredients-based approach including CT/NG and TV cartridges and consumables (collection kits, gloves, swabs), using Philani Ndiphile trial data and published and unpublished sources (Table 1).[42, 43] Personnel costs were estimated using published South African public sector salary scales. The cost of syndromic management was drawn from a previous South African study, including the cost of personnel, supplies, equipment and overheads.[44] Drug prices for syndromic management and the intervention arms were obtained from the South African Master Health Product List 2024[45] with utilisation in the intervention arms obtained from trial data.

Costs of infant complications due to preterm birth and low birthweight included potential hospitalisation during the first year of life.[46] Corresponding data on resource use, including pharmaceuticals, medical interventions, and average length of stay in intensive and high-dependency care for low birthweight infants and those with preterm birth-related respiratory complications were obtained from a 2024 South African study.[46] In the absence of neonatal intensive care unit cost data, cost per patient day was approximated using estimates for general intensive care units in South Africa.[47] Costs for high-dependency care and general ward admissions were based on the cost per patient day in a district hospital as reported in the South Africa District Health Barometer.[48] Further details on the calculation of infant hospitalisation costs are provided in Supplementary table 3. Preterm birth and low birthweight complication rates and associated hospitalisation rates and duration of stay are from published literature (Table 1).[33, 46, 49, 50]

### Model outcomes

Incremental cost-effectiveness ratios (ICER) and incremental cost-utility ratios (ICUR) were calculated by dividing the incremental cost of each intervention arm by the outcomes averted in comparison to standard care syndromic management. Cost-effectiveness results are presented as ICERs: incremental cost per composite and component outcomes averted. These results are interpreted as the relative cost-effectiveness of the two screening strategies to standard care, given that no established cost-effectiveness thresholds exist for interpreting ICERs expressed in natural outcome units.

In the absence of a recognised cost per DALY threshold (cost-effectiveness threshold) for cost–utility analyses, we evaluated ICURs using two published DALY-based thresholds: an “optimistic” threshold of $4,702 per DALY averted[51] and a “conservative” threshold of $3,526 per DALY averted.[52] Both values were inflated from 2015 to 2024 USD prices using average Rand/USD exchange rates and published consumer price inflation indices.[25, 26]

### Sensitivity and threshold analyses

We evaluated the robustness of the cost-effectiveness and cost-utility model findings through comprehensive sensitivity analyses. Univariate analysis identified the most influential parameters. We varied cost parameters based on low and high estimates from published literature, or fixed ranges (±30%) where estimates were not available. Probability and DALY-related parameters were adjusted using 95% confidence intervals from trial data, published literature [44, 45, 49, 50, 53] (Table 1 and Supplementary Table 2), or fixed ranges (±30%).

Probabilistic sensitivity analysis employed Monte Carlo simulation over 10,000 iterations to assess uncertainty in cost-effectiveness and cost-utility, assigning appropriate probability distributions to parameters (gamma distributions for costs; beta or log-normal distributions for probabilities and DALY parameters). Uncertainty around ICERs and ICURs using cost-effectiveness planes and 95% uncertainty intervals derived from simulations are presented alongside the base-case results.

## RESULTS

### Base case

The mean total cost of syndromic management (standard care) was $3.74 (95% CI: $3.72–$3.75) per pregnancy, compared with $63.13 ($62.55–$63.73) for One-Time Screening (incremental cost of $59.39 ($58.83–$59.98)) and $94.83 ($93.87–$95.79) for Two-Time Screening (incremental cost of $91.09 ($90.14–$92.04)) (Table 2).

**Table 2:**
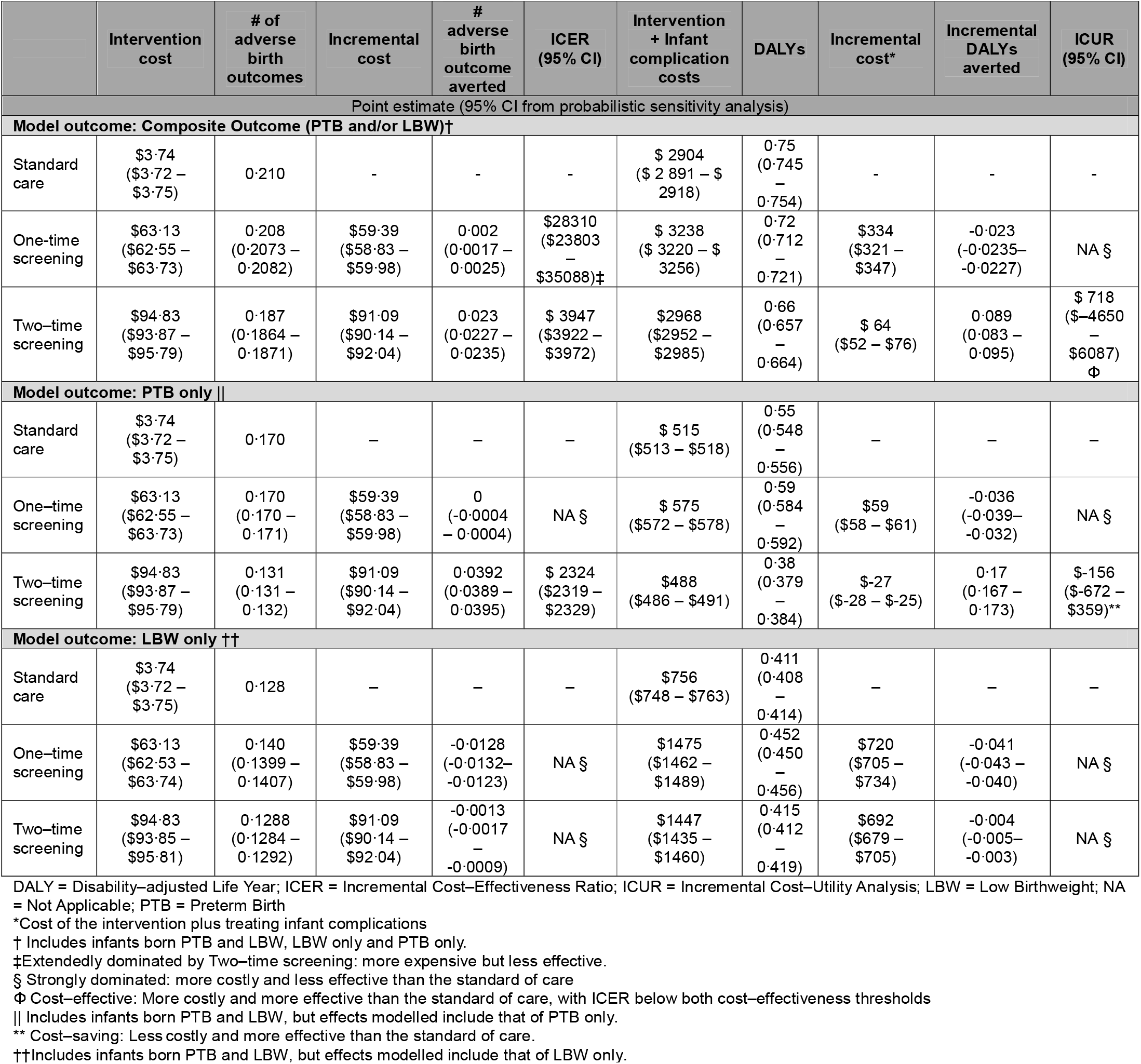
Base case and uncertainty analysis cost-effectiveness and cost-utility results; mean per pregnancy.

Intervention Cost-Effectiveness and -Utility for Composite Primary Outcome:

Compared with standard care, One-Time Screening averted 0.002 (0.0017–0.0025) composite outcomes per pregnancy (20 per 10,000 pregnancies), while Two-Time Screening averted 0.023 (0.0227–0.0235) cases (230 per 10,000 pregnancies). ICERs were $28,310 ($23,803 – $35,088) per composite outcome averted for One-Time and $3,947 ($3,922 – $3,972) for Two-Time Screening (Table 2). While both interventions improved health outcomes compared with standard care, and Two-Time Screening cost less to avert an additional composite outcome than One-Time Screening, both interventions cost more than standard care syndromic management.

Compared with standard care, One-Time Screening resulted in an additional 0.023 (0.0227 - 0.0235) DALYs per pregnancy (230 DALYs per 10,000 pregnancies), while Two-Time Screening averted 0.089 (0.083–0.095) DALYs (890 DALYs per 10,000 pregnancies). The mean total cost of standard care, hospitalisation and treatment related to preterm birth, low birthweight or both was $2,904 ($2,891–$2,918). In comparison, the mean total costs of One- and Two-Time Screening per pregnancy was $3,238 ($3,220–$3,256) and $2,968 ($2,952–$2,985), respectively, with incremental costs of $334 ($321–$347) and $64 ($52–$76), respectively (Table 2). Consequently, One-Time Screening was more costly and less effective than standard care (strongly dominated). Two-Time Screening, with an ICUR of $718 ($-4650–$6087) per DALY averted below both the optimistic and conservative cost-effectiveness thresholds, was cost-effective (Figure 2, Panel A).

**Figure 2:**
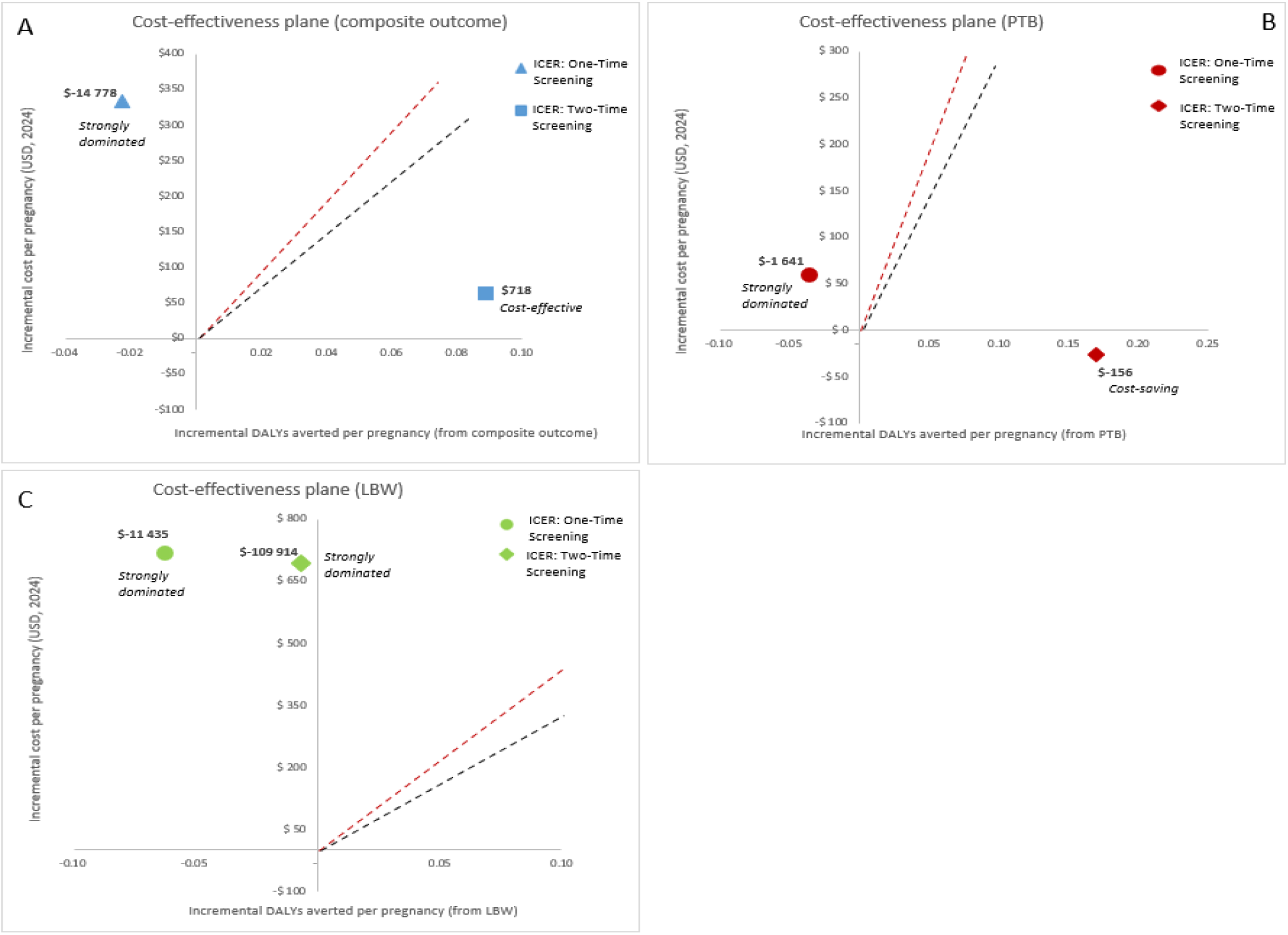
Cost-effectiveness plane (base case cost-utility results) Interpretation: Red dotted line represents the optimistic cost-effectiveness threshold ($4,702 per DALY averted); Black dotted line represents the conservative cost-effectiveness threshold ($3,526 per DALY averted) DALYs = disability-adjusted life years; ICER = incremental cost-effectiveness ratio; LBW = low birthweight; PTB = preterm birth.

Intervention Cost-Effectiveness and -Utility for Component Preterm Birth Outcome:

Compared with standard care, One-Time Screening did not avert any cases of preterm birth and was more costly (strongly dominated). In comparison, Two-Time Screening averted 0.0392 (0.0389–0.0395) cases of preterm birth per pregnancy (392 per 10,000 pregnancies), but at higher costs (ICER per preterm birth averted: $2,324 ($2,319–$2,329) (Table 2).

Compared with standard care, One-Time Screening resulted in an additional 0.036 (0.032–0.039) DALYs per pregnancy (360 DALYs per 10,000 pregnancy), while Two-Time Screening averted 0.17 (0.167–0.173) DALYs (1700 DALYs per 10,000 pregnancies). The total cost of screening, hospitalisation and treatment for preterm birth was $575 ($572–$578) for One-Time Screening, and $488 ($486–$491) for Two-Time Screening, resulting in an incremental cost of $59 ($58–$61) and savings of $27 ($25–$28) per pregnancy, respectively. One-Time Screening was more costly and less effective than standard care (strongly dominated) (Panel B). In comparison, Two-Time Screening yielded a negative incremental cost of $-156 ($-672–$359) per DALY averted, indicating lower cost and greater effectiveness than standard care (Figure 2, Panel B).

Intervention Cost-Effectiveness and -Utility for Component Low Birthweight Outcome:

One-Time Screening resulted in 0.0128 (0.0123 – 0.0132) additional cases of low birthweight per pregnancy (128 per 10,000 pregnancies) while Two-Time Screening resulted in 0.0013 (0.0009–0.0017) additional cases. Both strategies incurred incremental costs and are thus strongly dominated by standard of care (more costly and less effective) (Table 2).

One-Time Screening resulted in 0.041 (0.040–0.043) additional DALYs per pregnancy (410 per 10,000 pregnancies) while Two-Time Screening resulted in 0.004 (0.003–0.005) additional DALYs (40 per 10,000 pregnancies). Both strategies incurred incremental costs and were thus strongly dominated by standard care (more costly and less effective) (Table 2 and Figure 2, panel C).

### Univariate sensitivity analysis

Tornado diagrams for univariate sensitivity of key parameters in the cost-utility analysis are shown in Supplementary Figure 1. Corresponding analysis and tornado diagrams for the cost-effectiveness analysis is presented in Supplementary Figures 2–4.

Univariate Sensitivity Analysis for Composite Primary Outcome:

One-Time Screening was dominated in all univariate sensitivity analyses except at the most favourable RR for the composite outcome, where it became cost-saving (Supplementary Figure 1, Panel A). Two-Time screening (Supplementary Figure 1, Panel B) was cost-effective based on both the conservative and optimistic cost-effectiveness thresholds in most sensitivity analyses, becoming strongly dominated only at the least favourable (highest) RR for the composite outcome. At the lowest values modelled for the RRs of Two-Time Screening to reduce the composite outcome, preterm birth and the proportion of preterm birth in the composite outcome, two-time screening became cost-saving with ICURs ranging from $-3,648 to $-3,866 per DALY averted).

Univariate Sensitivity Analysis for Component Preterm Birth Outcome:

One-Time Screening (Supplementary Figure 1, Panel C), was strongly dominated (less effective and more costly) in all univariate sensitivity analyses, becoming cost-saving only when the RR for preterm birth was at its lowest plausible value. Two-Time screening was either cost-saving or cost-effective in all univariate sensitivity analyses (Supplementary Figure 1, Panel D).

Univariate Sensitivity Analysis for Component Low Birthweight Outcome:

One-Time (Supplementary Figure 1, Panel E) and Two-Time Screening (Supplementary Figure 1, Panel F) were strongly dominated in all univariate sensitivity analyses, becoming only cost-saving if the RR for preterm birth was at its lowest plausible value, yielding an incremental cost of $-870 and $-313 per DALY averted, respectively.

### Probabilistic sensitivity analysis

Results for the cost-utility analysis are shown in Figure 3; cost-effectiveness results are in Supplementary Figure 5.

**Figure 3:**
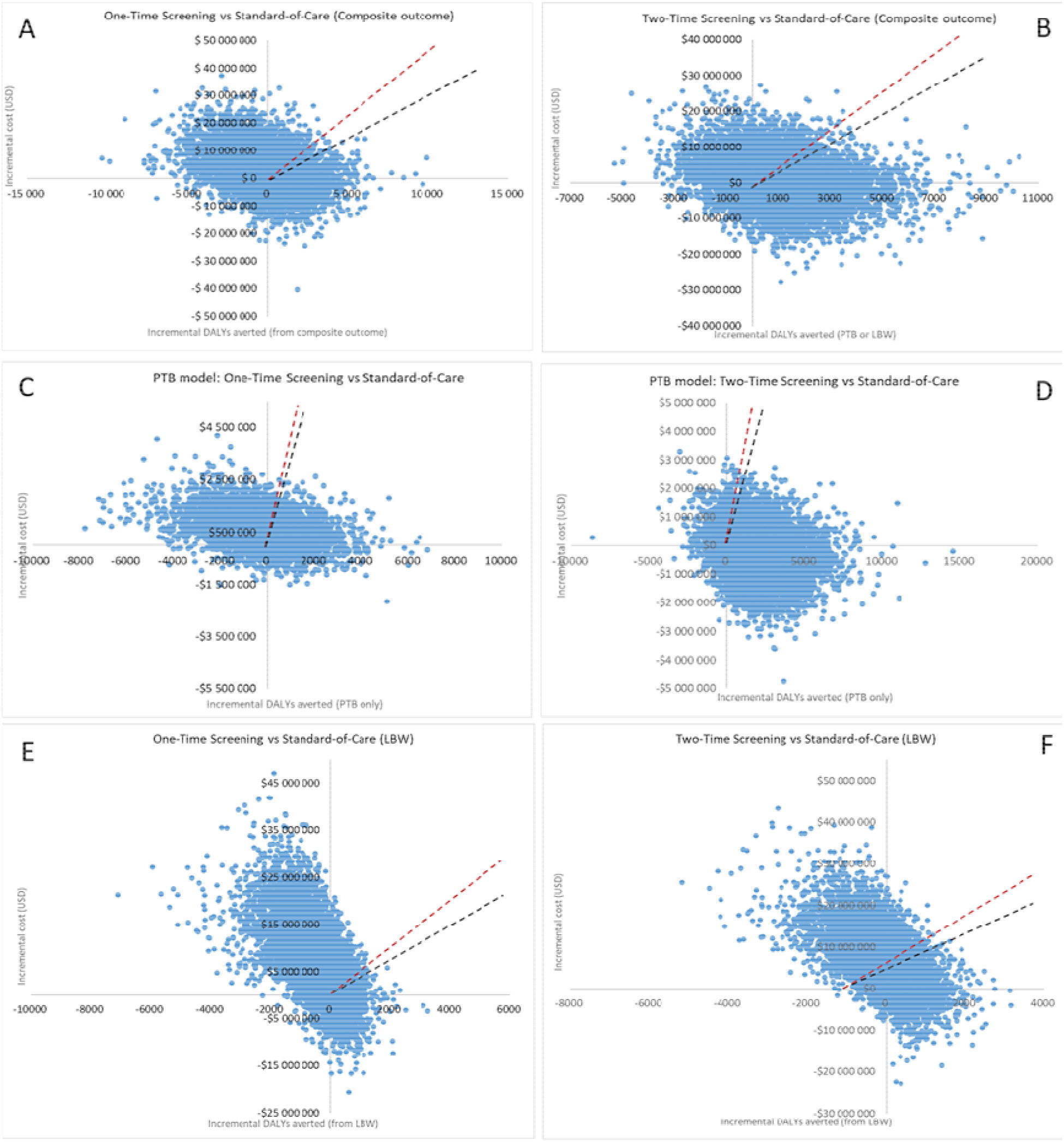
PSA cost-effectiveness planes (cost-utility analysis) Interpretation: Northeast quadrant: cost-effective when below respective cost-effectiveness thresholds (more costly and more effective than standard care taking resource constraints into account); Northwest quadrant: strongly dominated (more costly and less effective than standard care); Southwest quadrant: less costly and less effective, trade-offs required to determine if lower effectiveness is justified by the associated cost savings; Southeast: cost-saving (less costly and more effective than standard care) DALYs= disability-adjusted life years; ICER = incremental cost-effectiveness ratio LBW = low birthweight; PTB = preterm birth.

Probabilistic Sensitivity Analysis for Composite Primary Outcome:

For One-Time Screening (Figure 3, Panel A), 44% of iterations were strongly dominated (northwest quadrant), 27% were potentially cost-effective (more effective and more costly; northeast). Of the latter, only 13% and 11% of iterations were cost-effective at optimistic and conservative thresholds. 18% were cost-saving (southeast). For Two-Time Screening (Figure 3, Panel B), 36% of iterations were more effective and more costly (northeast) of which 20%–23% were cost-effective at optimistic and conservative thresholds, and 38% cost-saving (southeast). These values indicate a 58%–61% probability of being cost-effective or cost-saving, which gives low certainty in the base-case that Two-Time Screening could be cost-effective for averting the composite outcome.

Probabilistic Sensitivity Analysis for Component Pre-term Birth Outcome:

For One-Time screening (Figure 3, Panel C), 53% of iterations were dominated (northwest), 29% more effective and costly (northeast) of which 25–26% were cost-effective at optimistic and conservative thresholds, and 11% cost-saving (southeast). For Two-time screening (Figure 3, Panel D), 31% of iterations were more effective and costly (northeast), of which 30% were cost-effective, and 60% were cost-saving (southeast). These values give a 90% probability that Two-Time Screening is cost-effective or cost-saving for averting preterm birth, indicating high certainty for the base case finding.

Probabilistic Sensitivity Analysis for Component Low Birth Weight Outcome:

For One-Time screening (Figure 3, Panel E), 71% of iterations were dominated (northwest), 16% more effective and costly (northeast) of which only 2%–3% were cost-effective under optimistic and conservative cost-effectiveness thresholds, and 9% cost-saving (southeast). For Two-Time screening (Figure 3, Panel F), 50% were dominated (northwest), 37% more effective and costly (northeast) of which only and 6%–7% were cost-effective under optimistic and conservative thresholds, and 11% were cost-saving (southeast). There is thus a 11%–12% and 17%–18% chance, respectively of One-Time or Two-Time Screening being cost-effective or cost-saving, indicating high certainty that neither strategy is cost-effective for low birthweight.

## DISCUSSION

In this study of the cost effectiveness of STI screening during pregnancy to reduce adverse birth outcomes, we report three key findings. First, molecular screening for *C. trachomatis, N. gonorrhoeae,* and *T. vaginalis* was not conclusively cost-effective for reducing the composite primary outcome of preterm birth and/or low birthweight compared to syndromic management. Second, two-time molecular screening was cost saving for reducing the secondary outcome, preterm birth, compared to syndromic management. Third, molecular screening was not cost-effective for reducing the secondary outcome of low birthweight compared to syndromic management. Together, these findings suggest that screening for *C. trachomatis, N. gonorrhoeae*, and *T. vaginalis* twice during pregnancy could reduce preterm birth while generating net savings to the health system. However, further evidence is required on clinical effectiveness on this secondary outcome.

Importantly, these findings add to the growing, yet still limited, body of economic evidence on antenatal screening for *C. trachomatis, N. gonorrhoeae,* and *T. vaginalis* in Sub-Saharan Africa, some of which considered upstream STI outcomes only. [21, 54] Our analysis goes beyond STI outcomes by evaluating the effects on *N. gonorrhoeae,* and *T. vaginalis* alongside *C. trachomatis* and incorporating downstream outcomes (adverse birth outcomes and DALYs).

For the composite outcome, the base case analysis indicated that two-time screening might be cost-effective (ICUR: US$718 per DALY averted) but uncertainty around this estimate was substantial based on probabilistic sensitivity analysis (∼61% certainty). This uncertainty is consistent with, and largely driven by, the non-significant reduction in the composite outcome in the Philani trial (RR; 0.89 (0.72–1.09)). Consequently, although two-time screening may represent a potentially cost-effective strategy based on point estimates, the evidence is insufficient to conclude this definitively. Our findings partially align with a Botswanan study which projected that GeneXpert screening for *C. trachomatis and N. gonorrhoeae* twice during pregnancy would be cost-effective for reducing preterm and/or low birthweight (ICUR: US$581 per DALY averted). However, similar to our findings, probabilistic sensitivity analysis showed substantial uncertainty, suggesting limited confidence in this finding.[22] The evidence on clinical effectiveness also remains mixed. Although modelling South Africa suggests screening for *C. trachomatis, N. gonorrhoeae,* and *T. vaginalis* during pregnancy can reduce both adverse birth outcomes[55], the WANTAIM trial in Papua New Guinea[56] found no reduction in preterm birth or low birthweight with three rounds of screening and treatment compared with syndromic management. These findings concur with the evidence from the Philani trial that uncertainty remains around the effectiveness for the primary composite, emphasizing the need for caution when interpreting these results.

For the secondary outcome of preterm birth alone, two-time screening was cost-saving, with US$156 saved for each healthy year of life gained; a substantial portion of South Africa’s 2023 per capita health expenditure (US$537)[57]. Savings were driven primarily by reductions in neonatal hospitalisation and newborn care costs and remained robust in probabilistic sensitivity analyses, with all simulations remaining either cost-saving or cost-effective. Longer-term economic benefits of preventing preterm birth are likely to be even greater, given the downstream developmental and health consequences of prematurity that were not fully captured. Importantly, as molecular platforms become cheaper, or rapid point-of-care tests become more widely available, the economic attractiveness of antenatal STI screening may improve further. Notably, no previous studies evaluated the effectiveness of cost-effectiveness of antenatal STI screening considering preterm birth alone, and because this was a secondary outcome in the Philani trial, additional evidence is needed to confirm clinical effectiveness.

Neither One-Time nor Two-Time screening was cost-effective for preventing low birthweight. This finding contrasts with a modelling study from Botswana, which estimated that antenatal *C. trachomatis and N. gonorrhoeae* screening with GeneXpert would be cost-effective for reducing low birthweight.[58] The difference in findings is mainly explained by modelling assumptions around the effectiveness of screening and is further distinguished by our inclusion of *T. vaginalis*.

This study has several limitations. The analysis was conducted from a provider perspective, excluding societal costs, such as client out-of-pocket expenses or productivity losses. Costs for maternal complications and preterm labour management were excluded. The time horizon was restricted to the first year of life, capturing early outcomes and hospitalisation but excluding longer-term disability and care, which likely makes estimates conservative; however, data limitations precluded lifetime modelling. Effectiveness and cost inputs are derived from an Eastern Cape-based trial, with other parameters, like disability weights, from literature, which may limit generalisability. Uncertainty in the Global Burden of Disease estimates, particularly the distinction between preterm birth and low birthweight and the availability of recent disability weights, adds uncertainty. However, given the short modelling horizon, these assumptions have minimal impact, as the magnitude of DALY estimates are largely driven by YLL. Finally, gaps in local cost and epidemiological data required use of secondary sources rather than trial-derived estimates, contributing additional uncertainty despite sensitivity analyses.

## CONCLUSION

Screening for *C. trachomatis, N. gonorrhoeae*, and *T. vaginalis* at the first antenatal visit and again during the third trimester is not conclusively cost-effective for preventing the composite outcome of preterm birth and/or low birthweight, nor for preventing low birthweight alone. However, if preterm birth is the only adverse outcome prevented by antenatal STI screening, Two-Time Screening in pregnancy has a high likelihood of being cost saving. These findings suggest that repeat antenatal STI screening could reduce preterm births while lowering overall health-system costs in high-burden settings. However, further evidence on clinical effectiveness is needed to inform decision on routine implementation.

## Supporting information

Consolidated Health Economic Evaluation Reporting Standards

## Data Availability

Data available on reasonable request from the corresponding author.

## LIST OF ABBREVIATIONS

ANC: Antenatal Care
CHEERS: Consolidated Health Economic Evaluation Reporting Standards
CLD: Chronic Lung Disease
CPAP: Continuous Positive Airway Pressure
CT: Chlamydia trachomatis
DALY: Disability-Adjusted Life Year
ICER: Incremental Cost-Effectiveness Ratio
ICUR: Incremental Cost-Utility Ratio
LBW: Low Birthweight
NA: Not Applicable
NG: Neisseria gonorrhoeae
NICU: Neonatal Intensive Care Unit
PTB: Preterm Birth
PoC: Point-of-Care
PSA: Probabilistic Sensitivity Analysis
RDS: Respiratory Distress Syndrome
RR: Relative Risk
SM: Syndromic Management
STI: Sexually Transmitted Infection
TOC: Test-of-Cure
TV: Trichomonas vaginalis
USD: United States Dollar
WHO: World Health Organization
YLD: Years Lived with Disability
YLL: Years of Life Lost

## DECLARATIONS

### Ethics approval and consent to participate

Ethical approval for this study was granted by the University of Cape Town Human Research Ethics Committee (Ref: 676/2019). The study was conducted in accordance with the ethical principles outlined in the Declaration of Helsinki, and all applicable national and institutional guidelines for research involving human participants were followed. All participants in the Philani Ndiphile trial provided written informed consent, which was obtained and documented in accordance with the approved study protocol.

### Consent for publication

Not applicable

### Availability of data and materials

Data available on reasonable request from the corresponding author.

### Competing interests

The authors declare that they have no competing interests

### Funding

This work was supported by U.S. National Institute of Health (R01AI149339); The funder of the study had no role in the study design, data collection, data analysis, data interpretation, or writing of the report.

### Author contributors

Study conceptualisation: AMM, SC, JK, SC, and ESi; Funding acquisition: AMM and JK; Data curation: CB, MM and FM; Analysis: ESm; Study supervision and data validation: ESi and CB

Writing original draft of the manuscript: ESm; Review and editing the manuscript: CB, AMM, MM, FM, NL, AO, RP, JK, SC and ESi. All authors read and approved the final version of the manuscript, had full access to all the data, and are responsible for the decision to submit for publication.

## Acknowledgements

Not applicable

## Notes

### Competing Interest Statement

The authors have declared no competing interest.

### Author Declarations

Ethical approval for this study was granted by the University of Cape Town Human Research Ethics Committee (Ref: 676/2019). The study was conducted in accordance with the ethical principles outlined in the Declaration of Helsinki, and all applicable national and institutional guidelines for research involving human participants were followed.

