## Supplementary for "Cost-effectiveness and cost-utility of antenatal sexually transmitted infection screening to reduce preterm birth and low birthweight in South Africa"

Supplementary Materials

*Supplementary figure and tables*

Supplementary Table 1: PTB, LBW and overlapping outcomes in the trial

| **Birth outcome** | **One-time screening** | **Two-time screening** | **Standard of care** |
| --- | --- | --- | --- |
| Total LBW or PTB (% of live births) | 22·9% | 20·6% | 23·2% |
| Total LBW (% of live births) | 14·1% | 12·9% | 12·8% |
| *LBW and not preterm (% of LBW)* | *28·4%* | *47·0%* | *34·1%* |
| *LBW and preterm (% of LBW)* | *71·6%* | *53·0%* | *65·9%* |
| Total PTB (% of live births) | 18·9% | 14·5% | 18·8% |
| *Preterm and not LBW (% of PTB)* | *45·8%* | *52·7%* | *55·4%* |
| *LBW and preterm (% of LBW)* | *54·2%* | *47·3%* | *44·6%* |
| Total LBW and PTB (% of live births) | 20·3% | 13·7% | 16·8% |
| Total LBW and PTB (% of all PTB or LBW) | 44·6% | 32·0% | 35·4% |
| **% Preterm births (of all live births)** | **One-time screening** | **Two-time screening** | **Standard of care** |
| Total preterm births | 18·8% | 14·5% | 18·8% |
| Extremely Preterm (< 28 weeks) | 0·8% | 0·3% | 0·5% |
| Very Preterm (28 to <32 weeks) | 2·9% | 1·7% | 2·8% |
| Moderately Preterm (32 to < 34weeks) | 2·6% | 1·9% | 2·5% |
| Late Preterm (34 to <37 weeks) | 12.7% | 10.6% | 13.1% |

*Source: Medina-Marino et al. (2026)^1^*

Supplementary Table 2: Additional cost-effectiveness and cost-utility model inputs, probabilities and DALY parameters

|  | **Model comparator** | | |  |  |
| --- | --- | --- | --- | --- | --- |
|  | **Syndromic management** | **One-Time Screening** | **Two-time Screening** | **Distribution for PSA** | **Source** |
|  | **Point estimate (low and high) value** | | |  |  |
| **Baseline**  **(enrolment )** |  |  |  |  |  |
| % women with VDS at baseline | 15.0 (10.5-19.5) ^a^ | NA | NA | Beta | ^1^ |
| % VDS treated baseline | 70·8  (49·6-92·0)* | NA | NA | Beta | ^1^ |
| % VDS treated at week 30-34 visit | 63·6  (44·5-82·7)* | 60·9  (42·7-79·2)* | NA | Beta | ^1^ |
| % women with CT at baseline | NA | 14.9 (10.4-19.3) | 14.5 (10.1-18.8) | Beta | ^1^ |
| % women tested positive for CT that are treated at baseline | NA | 94·6 (66·3-100·0)^a^ | 93·5 (65·4-100·0)^a^ | Beta | ^1^ |
| % women with NG at baseline | NA | 4.9 (3.4-6.4) | 4.9 (3.4 -6.3) | Beta | ^1^ |
| % women tested positive for NG that are treated at baseline | NA | 83·8 (58·6-100·0)^a^ | 94·4 (66·1-100·0)^a^ | Beta | ^1^ |
| % women with TV at baseline | NA | 10.7 (7.5-14.0) | 9.6 (6.7-12.5) | Beta | ^1^ |
| % women tested positive for TV that are treated at baseline | NA | 95·1 (66·5-100·0)^a^ | 94·4 (66·1-100·0)^a^ | Beta | ^1^ |
| % of women treated for CT treated with Azithromycin 1g stat dos at baseline | 98·7 | 99·1 | 100·0 | Not varied | ^1^ |
| % of women treated for CT treated with Azithromycin 2g stat dos at baseline * | 1·3 | 0·9 | 0·0 | Not varied | ^1^ |
| % of women treated for NG treated with Ceftriaxone 250mg IM at baseline * | 92·1 | 54·8 | 61·8 | Not varied | ^1^ |
| % of women treated for NG treated with Ceftriaxone 1g IM at baseline * | 5·3 | 6·5 | 20·6 | Not varied | ^1^ |
| % of women treated for NG treated with Ceftriaxone 500mg IM at baseline * | 3·9 | 38·7 | 17·6 | Not varied | ^1^ |
| % of women treated for TV treated with Metronidazole 400mg bdweek at baseline * | 94·7 | 93·5 | 95·5 | Not varied | ^1^ |
| % of women treated for TV treated with Metronidazole 2g stat dos at baseline * | 1·3 | 6·5 | 4·5 | Not varied | ^1^ |
| **Test-of-cure (TOC)** |  |  |  |  |  |
| % women tested positive for CT returning for TOC | NA | 14.3 (10.0-18.6)^a^ | NA | Beta | ^1^ |
| % women tested positive for NG returning for TOC | NA | 58.0 (40.6-75.4)^a^ | NA | Beta | ^1^ |
| % women tested positive for TV returning for TOC | NA | 51.4 (35.9-66.8)^a^ | NA | Beta | ^1^ |
| % of women returning that are tested: CT/NG | NA | 49.1 (34.4-63.8) ^a^ | NA | Beta | ^1^ |
| % of women returning that are tested: TV | NA | 22.2 (15.6-28.9) ^a^ | NA | Beta | ^1^ |
| % of women returning that are tested: CT/NG & TV | NA | 28.7 (20.1-37.3) ^a^ | NA | Beta | ^1^ |
| % women with CT at TOC | NA | 10.7 (7.5-13.9) | NA | Beta | ^1^ |
| % women tested positive for CT treated | NA | 66.7 | NA | Not varied | ^1^ |
| % women with NG at TOC | NA | 2.6 (1.8-3.3) | NA | Beta | ^1^ |
| % women tested positive for NG treated | NA | 50.0 | NA | Not varied | ^1^ |
| % women with TV at TOC | NA | 20.0 (14.0-26.0) | NA | Beta | ^1^ |
| % women tested positive for TV treated | NA | 100.0 | NA | Not varied | ^1^ |
| % of women treated for CT treated with Azithromycin 1g stat dos | NA | 100.0 | NA | Not varied | ^1^ |
| % of women treated for CT treated with Azithromycin 2g stat dos | NA | 0.0 | NA | Not varied | ^1^ |
| % of women treated for NG treated with Ceftriaxone 250mg IM | NA | 100.0 | NA | Not varied | ^1^ |
| % of women treated for NG treated with Ceftriaxone 1g IM | NA | 0.0 | NA | Not varied | ^1^ |
| % of women treated for NG treated with Ceftriaxone 500mg IM | NA | 0.0 | NA | Not varied | ^1^ |
| % of women treated for TV treated with Metronidazole 400mg bdweek | NA | 90.9 | NA | Not varied | ^1^ |
| % of women treated for TV treated with Metronidazole 2g stat dos | NA | 9.1 | NA | Not varied | ^1^ |
| ***Week 30-34 ANC visit*** |  |  |  |  |  |
| % enrolled participants returning for week ~32 visit | 67·5 (47·3-87·8) ^a^ | 64.3 (45.0-83.6)^a^ | 69.4 (48.6-90.2) | Beta | ^1^ |
| % of women returning with VDS | 12.9 (9.1-16.8) | 13.2 (9.2-17.2) | NA | Beta | ^1^ |
| % of women returning that are tested: CT/NG only | NA | NA | 0.2 (0.1-0.3) ^a^ | Beta | ^1^ |
| % of women returning that are tested: CT/NG & TV | NA | NA | 97.3 (68.1-100) ^a^ | Beta | ^1^ |
| % women with CT | NA | NA | 4.8 (3.4-6.3) | Beta | ^1^ |
| % women tested positive for CT treated | NA | NA | 87.5 (61.3-100.0) ^a^ | Beta | ^1^ |
| % women with NG | NA | NA | 2.6 (1.8-3.4) | Beta | ^1^ |
| % women tested positive for NG treated | NA | NA | 84.6 (59.2-100.0) | Beta | ^1^ |
| % women with TV | NA | NA | 2.8 (2.0-3.7) | Beta | ^1^ |
| % women tested positive for TV treated | NA | NA | 92.9 (65.0-100.0) | Beta | ^1^ |
| % of women treated for CT treated with Azithromycin 1g stat dos * | 92·9 | 92.3 | 100.0 | Not varied | ^1^ |
| % of women treated for CT treated with Azithromycin 2g stat dos * | 2·4 | 2.6 | 0.0 | Not varied | ^1^ |
| % of women treated for NG treated with Ceftriaxone 250mg IM * | 95·2 | 87.2 | 18.2 | Not varied | ^1^ |
| % of women treated for NG treated with Ceftriaxone 1g IM * | 0 | 0.0 | 45.5 | Not varied | ^1^ |
| % of women treated for NG treated with Ceftriaxone 500mg IM * | 0 | 5.1 | 36.4 | Not varied | ^1^ |
| % of women treated for TV treated with Metronidazole 400mg bdweek * | 4·8 | 12.8 | 33.3 | Not varied | ^1^ |
| % of women treated for TV treated with Metronidazole 2g stat dos * | 92·9 | 76.9 | 66.7 | Not varied | ^1^ |
| ***Birth outcomes*** |  |  |  |  |  |
| % of participants where either a live birth of pregnancy loss was recorded | 94·0 (65·8-100·0) ^a^ | 93.2 (65.3-100.0) ^a^ | 95.7 (67.0-100.0) ^a^ | Beta | ^1^ |
| % live births | 90·6 (63·4-100·0) ^a^ | 88.9 (62.2-100.0) ^a^ | 90.9 (63.6-100.0) ^a^ | Beta | ^1^ |
| % pregnancy loss | 9·4 (6·6-12·3) ^a^ | 11.1 (7.8-14.5) ^a^ | 9.1 (6.4-11.8)^a^ | Beta | ^1^ |
| % Total preterm births [of all live births] | 18·8 (13·2-24·5) ^a^ | 18.8 (13.2-24.5) ^a^ | 14.5 (10.1-18.8) ^a^ | Beta | ^1^ |
| % Extremely Preterm (< 28 weeks) [of all live births] | 0·5 (0·3-0·6) ^a^ | 0.8 (0.6-1.0) ^a^ | 0.3 (0.2-0.4) ^a^ | Beta | ^1^ |
| % Very Preterm (28 to <32 weeks) [of all live births] | 2·8 (2·0-3·6) ^a^ | 2.9 (2.0-3.7) ^a^ | 1.7 (1.2-2.2) ^a^ | Beta | ^1^ |
| % Moderately Preterm (32 to < 34weeks) [of all live births] | 2·5 (1·7-3·2) ^a^ | 2.6 (1.8-3.3) ^a^ | 1.9 (1.3-2.4) ^a^ | Beta | ^1^ |
| % Late Preterm (34 to <37 weeks) [of all live births] | 13·1 (9·1-17·0)^a^ | 12.6 (8.8-16.4) ^a^ | 10.6 (7.4-13.8) ^a^ | Beta | ^1^ |
| % born preterm (any) or LBW [of all live births] | 23·2 (16·2-30·1) ^a^ | 22.9 (16.1-29.8) ^a^ | 20.6 (14.4-26.8) ^a^ | Beta | ^1^ |
| % born LBW [of all live births] | 12·8 (8·9-16·6)^a^ | 14.0 (9.8-18.2) ^a^ | 12.9 (9.0-16.7) ^a^ | Beta | ^1^ |
| % preterm babies, not LBW | 23·1 (16·2-30·1) ^a^ | 45.8 (32.0-59.5)^a^ | 52.7 (36.9-68.5) ^a^ | Beta | ^1^ |
| % LBW babies, not preterm | 65·9 (46·1-85·6) ^a^ | 28.4 (19.9-36.9)^a^ | 47.0 (32.9-61.1) ^a^ | Beta | ^1^ |
| ***DALY inputs*** |  |  |  |  |  |
| *Early neonatal mortality following term birth* | 0·012 (0·009-0·016) ^b^ | | | Beta | **^2^** |
| *Late neonatal mortality following term birth* | 0·003 (0·002 -0·004) ^b^ | | | Beta | **^2^** |
| *Post-neonatal mortality following term birth* | 0·008 (0·006-0·011) ^b^ | | | Beta | **^2^** |
| Early Neonatal mortality (death within first 6 days of life) from any cause | 0·019 (0·013-0·024)^a^ | | | Beta | **^2^** |
| Late Neonatal mortality (death within 8-27 days of life) from any cause | 0·004 (0·003-0·005)^a^ | | | Beta | **^2^** |
| Post- Neonatal mortality (death within 1-11 months life) from any cause | 0·009 (0·006-0·011)^a^ | | | Beta | **^2^** |
| Early Neonatal mortality (death within first 6 days of life) from PTB | 0·006 (0·005-0·008)^a^ | | | Beta | **^2^** |
| Late Neonatal mortality (death wihtin 8-27 days of life) from PTB | 0·001 (0·0007-0·0012)^a^ | | | Beta | **^2^** |
| Post- Neonatal mortality (death wihtin 1-11 months life) from PTB | 0·0003 (0·0002-0·0003)^a^ | | | Beta | **^2^** |

Note: CT= Chlamydia trachomatis; NG= Neisseria gonorrhoeae; TV= Trichomonas vaginalis; stat dos = once-off dose; bdweek = twice a day for 7 days; IM = intramuscular injection; Probability-based and utility-based parameters were adjusted using either 95% confidence intervals from empirical trial data, published literature, or fixed ranges (±30%) where these were not available.

^a^ Upper/lower bound defined as a 30% increase/decrease from the point estimate

^b^ Upper/lower bound obtained from published literature

^*^ As targeted treatment of detected pathogen or as part of syndromic management regimen

**Supplementary Table 3: Respiratory distress syndrome (RDS) and chronic lung disease (CLD) hospitalization costs**

|  | **Unit cost** | **Resource utilization** | **Total cost** | **Source** |
| --- | --- | --- | --- | --- |
| **RDS-related hospitalization** |  |  |  |  |
| Hospital stay (days) | $ 214·46 | 14 | $ 3 002·39 | Cost^3^; Resource utilization^4^ |
| CPAP (days) | $ 534·29 | 1 | $ 534·29 | ^4^ |
| Nasal prong oxygen (days) | $ 36·47 | 14 | $ 510·59 | ^4^ |
| Surfactant (doses) | $ 126·05 | 2 | $ 252·09 | ^4^ |
| **Total cost per episode** |  |  | **$ 4 299·36** |  |
| **Average cost per hospital day** |  |  | **$ 307·10** |  |
| **Average cost per hospital day (excluding hospital stay cost)** |  |  | **$ 92·64** |  |
| **CLD-related hospitalization** |  |  |  |  |
| Hospital stay (days) | $ 214·46 | 28 | $ 6 004·77 | Cost^3^; Resource utilization^4^ |
| Lasix 0.5-1mg/kg/dose 12 hourly | $ 36·47 | 9 | $ 328·23 | ^4^ |
| Spironolatone 1mg/kg/dose 12hourly | $ 19·10 | 9 | $ 171·94 | ^4^ |
| Salbutamol – 0.1-0.5 mg/kg 2-6 hourly | $ 3·07 | 1 | $ 3·07 | ^4^ |
| Dexamethasone 0.05 mg/kg/dose daily X 3 days, 0.025mg/kg/dose daily X 3 days | $ 14·26 | 6 | $ 85·56 | ^4^ |
| **Total cost per episode** |  |  | **$ 6 593·57** |  |
| **Average cost per hospital day** |  |  | **$ 235·48** |  |
| **Average cost per hospital day (excluding hospital stay cost)** |  |  | **$ 21·03** |  |

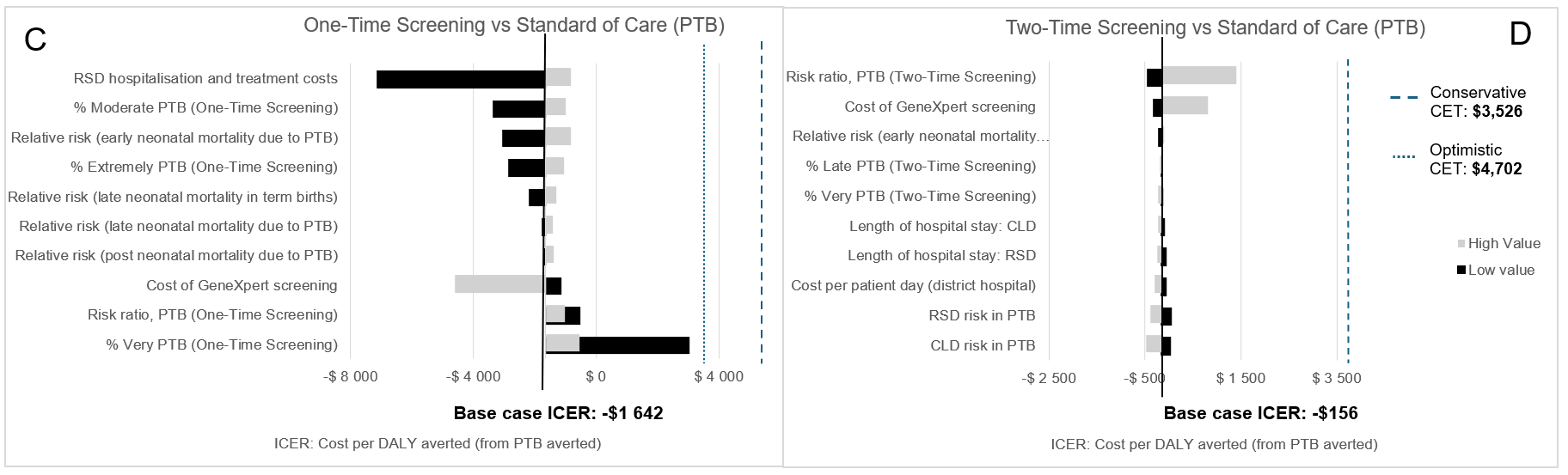

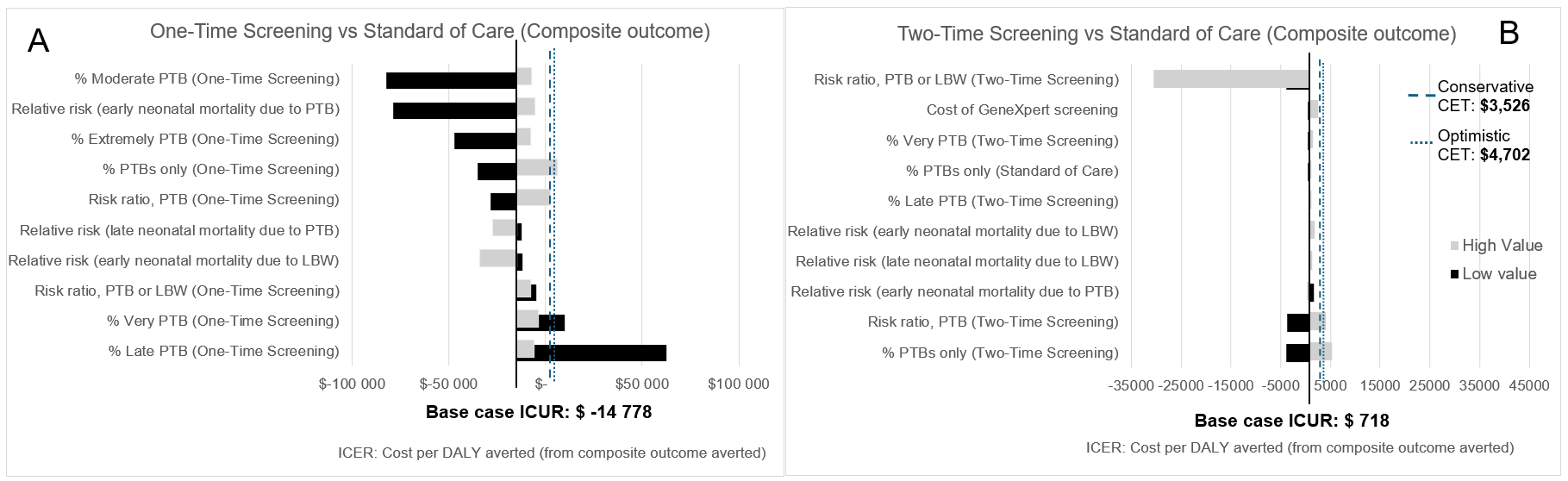

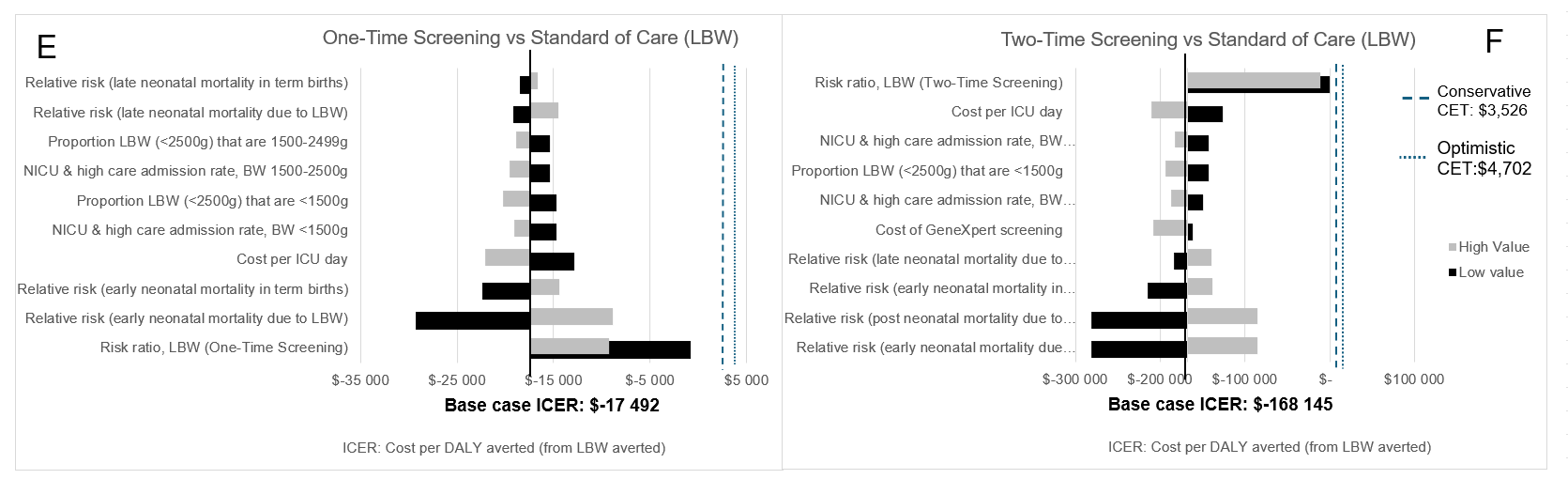

CET = cost-effectiveness threshold; DALYs = disability-adjusted life years; ICER = incremental cost-effectiveness ratio; NICU = neonatal intensive care unit; LBW = low birthweight; PTB = preterm birth.

Supplementary figure 1: Univariate sensitivity analysis of 10 most influential parameters (cost-utility analysis)

Supplementary figure 2: Univariate sensitivity analysis of 10 most influential parameters (cost-effectiveness analysis): One-Time Screening and Two-Time Screening vs Standard of Care (Composite outcome)

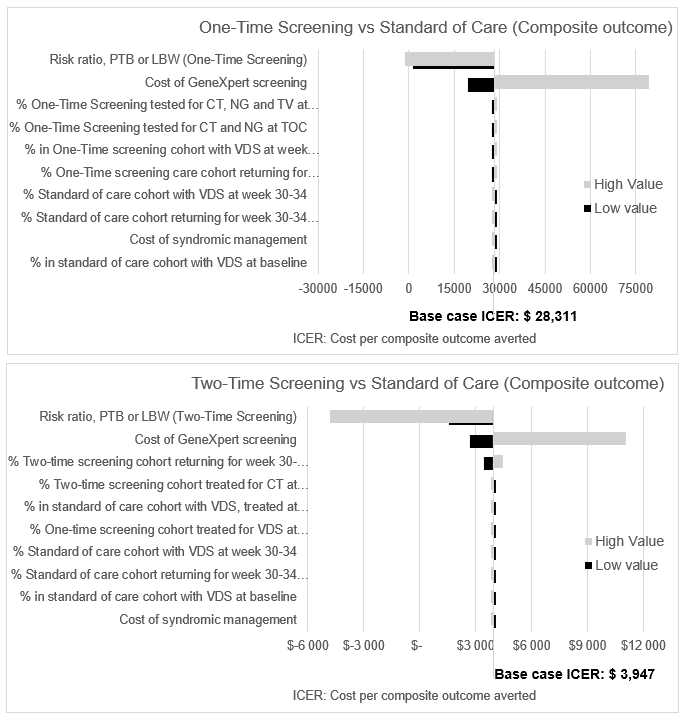

The relative cost-effectiveness of both One- and Two-Time Screening was most sensitive to the relative risk of the composite outcome and the cost of testing (currently GeneXpert). However, Two-Time Screening remained the most cost-effective in all one-way sensitivity analyses, except when the relative risk for reducing the composite outcome was at its least favourable extreme compared with standard care.

Negative incremental cost-effectiveness ratios (ICERs) for One-Time and Two-Time Screening all fall within the northwest quadrant of the cost-effectiveness plane (strongly dominated; more costly and less effective than the standard of care).

Positive ICERs for One-Time and Two-Time Screening all fall within the northeast quadrant of the cost-effectiveness plane (potentially cost-effective: more costly and more effective than the standard of care).

Supplementary figure 3: Univariate sensitivity analysis of 10 most influential parameters on cost-effectiveness results: One-Time Screening vs Standard of Care (Preterm Birth)

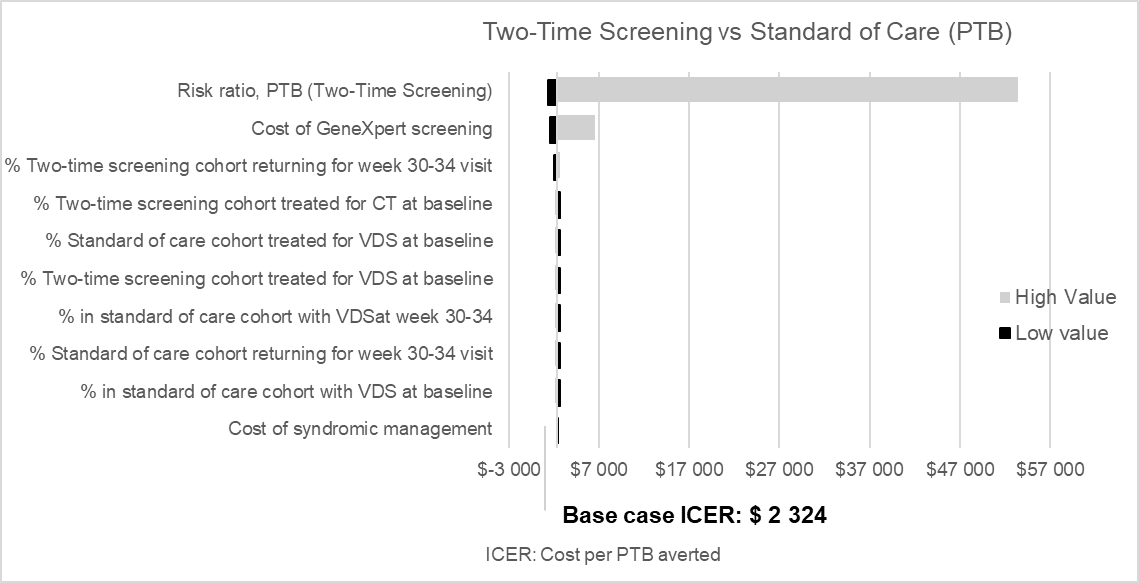

The cost-effectiveness of One-Time Screening was only affected by changes in the risk ratio for PTB and is subsequently not shown on a Tornado diagram.

Supplementary figure 4: Univariate sensitivity analysis of 10 most influential parameters on cost-effectiveness results: One-Time Screening and Two-Time Screening vs Standard of Care (Low Birthweight)

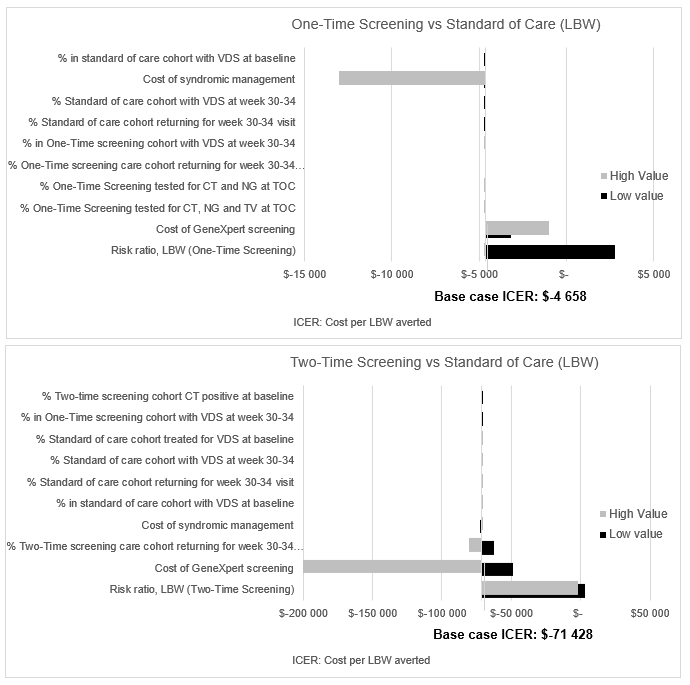

Both One-Time and Two-Time Screening remained dominated by standard care, except at the respective lowest relative risks for LBW, where either strategy could become potentially cost-effective (more costly but more effective than standard care).

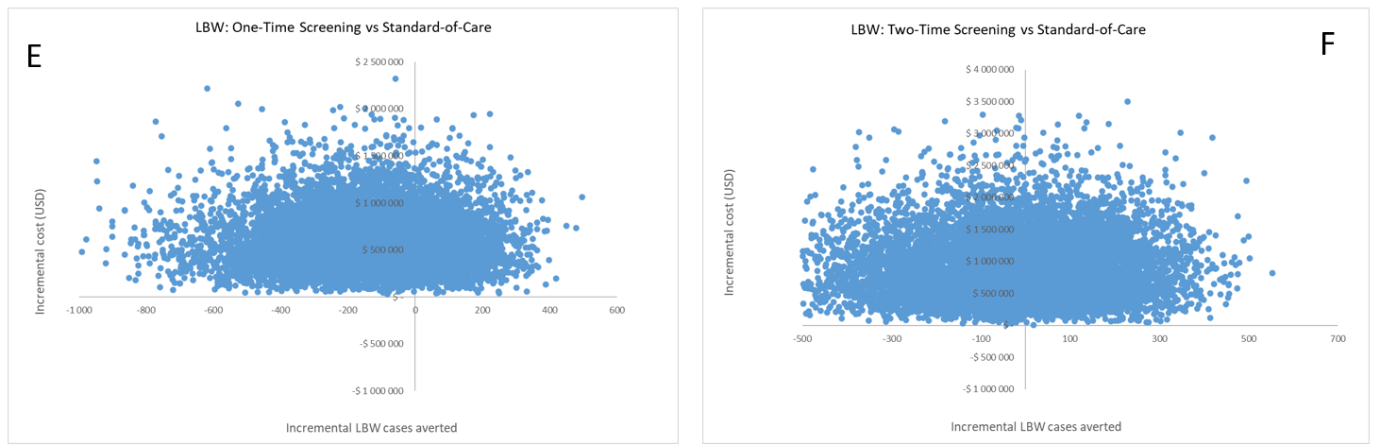

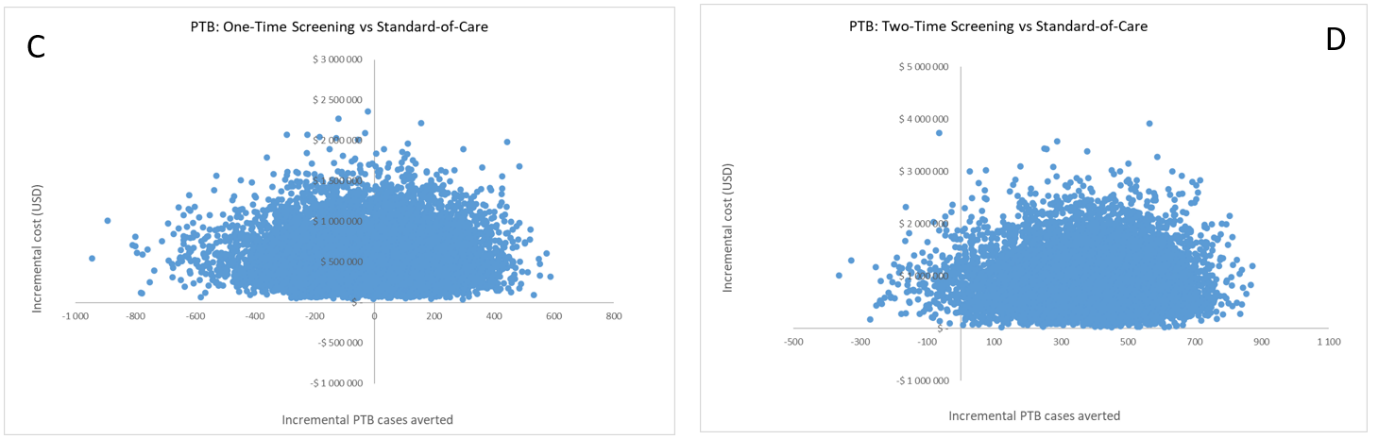
Supplementary figure 5: Probabilistic sensitivity analysis cost-effectiveness planes (cost-effectiveness analysis)

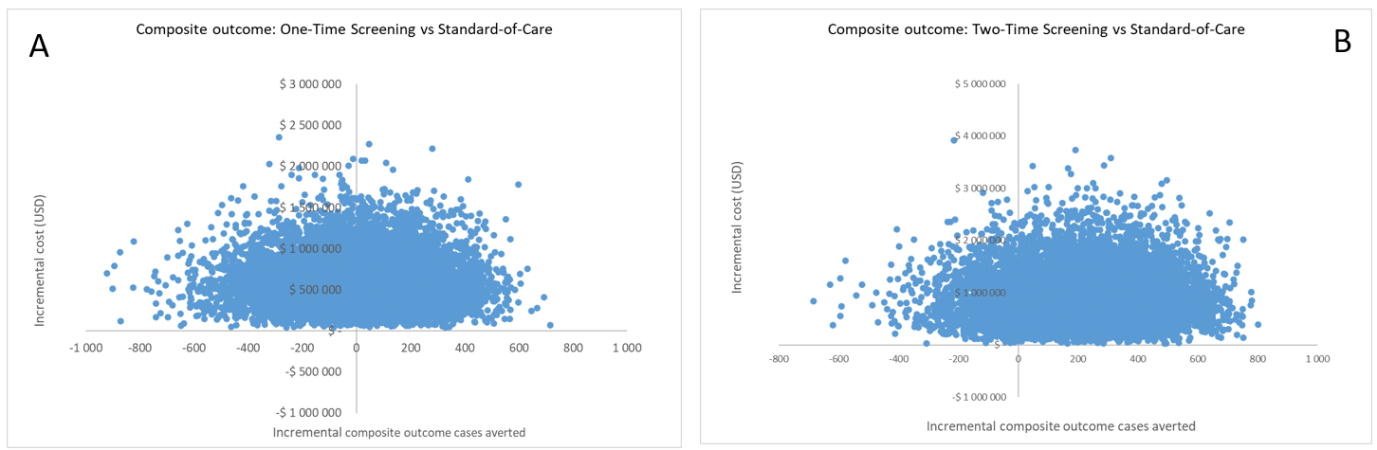

Composite outcome - Panel A and B: For One-Time Screening, Monte Carlo simulations are divided between the northeast quadrant (54%; more effective and more costly than standard care) and the northwest quadrant (46%; less effective and more costly) (panel A). For Two-Time Screening, 86% of Monte Carlo simulations fell in the northeast quadrant, and 14% in the northwest quadrant (panel B).

Preterm birth - Panel C and D: For One-Time Screening, Monte Carlo simulation are evenly divided between the northeast (50%; more effective and more costly) and northwest quadrants (50%; less effective and more costly) (panel C). For Two-Time Screening, 98% of simulations fall in the northeast quadrant, and 2% in the northwest quadrant (panel D).

Low birthweight - Panel E and F: For One-Time Screening, 75% of Monte Carlo simulations are in the northwest quadrant (less effective and more costly), 25% are in the northeast quadrant (more effective and more costly) (panel E). For Two-Time Screening, 52% of simulations are in the northwest quadrant (less effective and more costly), 48% are in the northeast quadrant (more effective and more costly) (panel F).
